# Using Neurocognitive Assessments to Predict and Differentiate Dementias: A Protocol for a Systematic Review

**DOI:** 10.1101/2023.01.10.23284251

**Authors:** Phuong Thuy Nguyen Ho, Christopher Li-Hsian Chen, Eric Josiah Tan

**Affiliations:** Memory, Ageing & Cognition Centre, National University Health System, Singapore; Department of Pharmacology, Yong Loo Lin School of Medicine, National University of Singapore, Singapore; Centre for Mental Health and Brain Sciences, School of Health Sciences, Swinburne University of Technology, Australia

## Abstract

Neurocognitive impairment is the primary feature of all types of dementia. This article presents the protocol for a systematic review of the available literature on the associations between cognitive task performance and various aspects of dementia prediction, specification and intervention outcomes.

## INTRODUCTION

Dementia is a highly prevalent disease, with age being one of the most stable predictors (1). As the average life expectancy increases globally, the prevalence of dementia also increases. It is estimated that over 75 million people will be diagnosed with dementia by 2030, most of whom will be living in low- or middle-income countries (2). In recent years, researchers have been focusing on different biomarkers to predict dementia and lessen the burden of this disease (3). While this field of research is up-and-coming, the high cost and inaccessibility of such markers can be an obstacle to clinical applications. An alternative potential marker that has emerged is neurocognitive testing, which can be carried out easily, is relatively more cost-effective and accessible.

Tracking the cognitive status of people with dementia through cognitive testing is important. Previous studies have found robust cognitive decline trajectories throughout the course of dementia (4). Furthermore, a randomized controlled trial in 2016 showed that baseline cognitive status can predict functional outcomes after intervention (5). In their 2007 review, Jacova et al. collated growing evidence for the utility of neuropsychological/cognitive assessments in relation to dementia prediction, differentiation between types of dementia, and progression of dementia from normal aging and mild cognitive impairment (MCI) (6) In 2017, Belleville also reviewed research findings on neuropsychological/cognitive measures predicting the progression from MCI to Alzheimer’s disease (AD) (7). Their meta-analysis found promising results for using neuropsychological assessments in predicting the progression of AD. Therefore, neuropsychological tests may be potentially helpful in predicting dementia as well as differentiating types of dementia.

Assessing a patient’s cognitive status can thus help health care professionals make sound decisions regarding treatment as well as interventions. In addition to synthesizing the current literature on the usage of neurocognitive tests on dementia prediction and differentiation, this review also aims to characterize the predictive values of neurocognitive assessments in predicting cognitive decline in dementia and intervention outcomes.

The research questions that will be addressed in this review are:

1. What neurocognitive measures can predict dementia onset or conversion?
2. What neurocognitive measures can differentiate subtypes of dementia?
3. What neurocognitive measures can predict the rate of decline of dementia?
4. What neurocognitive measures can predict intervention outcomes?

## METHODS

### Eligibility criteria

#### Population

Studies with adult human participants (at least 18 years old) who do not have chronic terminal illnesses, or other neurological (excluding stroke), genetic, and developmental disorders. Animal studies will be excluded.

#### Intervention

Observational studies or intervention studies that included at least one published neurocognitive test. Intervention studies must include baseline neurocognitive measures. Studies that investigated the combined effects of neurocognitive tests and other elements without studying individual effect of neurocognitive tests will be excluded.

#### Outcome

Four main outcomes will be assessed:

1. Dementia onset/conversion
2. Types of dementia
3. Cognitive decline in dementia
4. Functional outcomes of interventions

Only studies using formal DSM or ICD diagnoses of dementia, or supported by medical examination, will be included.

#### Study design

The current review will include observational as well as intervention studies. Case-reports, case-control studies, conference proceedings, reviews, meta-analyses and other grey literature will be excluded.

#### Further restrictions

Only studies published in peer-reviewed journals and written in English will be included. We will only include studies that employed objective neurocognitive tests.

### Information sources and search strategy

The search will be performed on three search engines: PubMed, Web of Science, and PsycINFO. The full search strategies for all three databases (adjusted slightly between them) are included in the Appendix. The search strategy will include three major themes: (1) neurocognitive tests, (2) dementia, (3) prediction and differentiation. The search for articles will be performed in January 2023 across the three databases. All papers up to this date will be included.

### Study records

#### Data management

Endnote will be utilized to store articles, remove duplicates, and retrieve full texts (8). A PRISMA flow diagram will be included in the review for better visualization of data selection and inclusion (9).

#### Selection process

The studies will first be screened based on the abstract by EJT and PTNH. Articles with abstracts that do not satisfy the inclusion criteria will be excluded. Afterwards, we will retrieve the full text of the remaining articles and further evaluate their eligibility. Any disagreements will be resolved within the authorship team. We will also screen the reference lists of the interim papers to identify further relevant studies.

#### Data collection process and data items

The two reviewers will independently extract the data items relating to study aims and methodology, sample characteristics, and primary findings pertaining to neurocognition from the included articles. The following data will be extracted: study background (authors, year of publication, country of dataset), sample characteristics (number of participants, age, percentage of female participants, study design and target population, neurocognitive tests used, diagnostic/prognostic outcome, statistical analysis and software used, any confounding variables reported, and main findings.

### Outcomes and prioritization

In this review, we will be looking at four specific outcomes: (1) dementia onset/conversion, (2) types of dementia, (3) cognitive decline in dementia, (4) outcomes of interventions. Dementia diagnosis will be the primary outcome due to the large number of available studies on this topic.

### Risk of bias assessment and study quality

We will perform the risk of bias assessment using the Joana Briggs Institute Critical Appraisal Tool for Systematic Reviews (https://jbi.global/critical-appraisal-tools) (10). The quality of the studies will be independently assessed by the two reviewers according to the Newcastle-Ottawa Assessment Scale (11). Any disagreements will be resolved between the authors.

### Data synthesis

The data will be synthesized in line with the aims and research questions previously outlined. The synthesis will highlight associations between neurocognition and key aspects of dementia in the adult population. Summaries of sample characteristics, study design, major findings and outcomes will also be detailed.

*This systematic review protocol was prepared in accordance with PRISMA-P guidelines (12).

## Data Availability

This systematic review protocol contains no new data.

## APPENDIX

### Search strategy for PubMed

*Concept #1: Dementia*

(alzheimer’s disease) OR (dementia)

*Concept #2: Neuropsychological tests*

(((cogniti*) OR (memory)) OR (neuropsychological test*))

*Concept #3: Prediction, progression, conversion*

(((((((predict*) OR (diagnos*)) OR (detect*)) OR (progress*)) OR (conversion)) OR (trajector*)) OR (Differentiat*)) OR (Distinguish)

*Concept #4: Exclusion of irrelevant terms*

(((((((((((((((((((((((((((((((((((((((((((((((((((((((((((((((((((((((((((((((((((((((((((Review) OR (Meta-analysis)) OR (huntington disease)) OR (Genetic disorder*)) OR (HIV)) OR (Developmental disorder*)) OR (Multiple Sclerosis)) OR (Psychosis)) OR (Schizophrenia)) OR (Tumor*)) OR (Autism)) OR (Epilepsy)) OR (Cerebral palsy)) OR (Amyotrophic Lateral Sclerosis)) OR (seizure*)) OR (infection*)) OR (Diabetes Mellitus)) OR (injury)) OR (rat)) OR (mouse)) OR (animal*)) OR (enzyme)) OR (Molecular)) OR (surgery)) OR (dog*)) OR (Alcoholism)) OR (chemical)) OR (neurochemical)) OR (Polymorphism)) OR (serum)) OR (lipid)) OR (protein)) OR (Pathogenesis)) OR (Epigenetic)) OR (receptor*)) OR (cell)) OR (Game*)) OR (food)) OR (damage*)) OR (glaucoma)) OR (Depression)) OR (eye*)) OR (optic)) OR (Caregiving)) OR (Caregiver*)) OR (Myelination)) OR (covid)) OR (glucose)) OR (gut))) OR (Inhibitor*)) OR (mutation*)) OR (cancer)) OR (apnea)) OR (Pollution)) OR (Amyloidosis)) OR (Case report*)) OR (Qualitative)) OR (arteriolosclerosis)) OR (virtual)) OR (urine)) OR (urinary)) OR (protocol*)) OR (syndrome)) OR (Cholesterol)) OR (Neurotransmitter*)) OR (Anticholinergic)) OR (glutamate)) OR (insulin)) OR (Postmenopausal)) OR (stroke*)) OR (delirium)) OR (Patient-centered)) OR (gait)) OR (Person-centered)) OR (fracture)) OR (case stud*)) OR (hygiene)) OR (anxiety)) OR (PTSD)) OR (ADHD)) OR (bipolar)) OR (Phospholipid*)) OR (phosphate)) OR (dental)) OR (tooth)) OR (chew)) OR (attitude*)) OR (Aggressi*)) OR (molecule*)) OR (obesity)) OR (insomnia)

#### Final search string

Search title/abstract: **#1** AND **#2** AND **#3** + search all fields NOT **#4**

### Search strategy for Web of Science

*Concept #1: Dementia*

(((TI=(dementia)) OR AB=(dementia)) OR AB=(alzheimer)) OR TI=(alzheimer)

*Concept #2: Neuropsychological tests*

(((((TI=(memory)) OR TI=(cogniti*)) OR TI=(neuropsychological test*)) OR

AB=(memory)) OR AB=(cogniti*)) OR AB=(neuropsychological test*)

*Concept #3: Prediction, progression, conversion*

(((((((((((((((TI=(diagnos*)) OR TI=(predict*)) OR TI=(detect*)) OR TI=(conversion)) OR

TI=(progress*)) OR TI=(distinguish)) OR TI=(Differentiat*)) OR TI=(trajector*)) OR

AB=(diagnos*)) OR AB=(predict*)) OR AB=(detect*)) OR AB=(conversion)) OR

AB=(progress*)) OR AB=(distinguish)) OR AB=(Differentiat*)) AND AB=(trajector*)

*Concept #5: Exclusion of irrelevant terms*

((((((((((((((((((((((((((((((((((((#4) NOT ALL=(review)) NOT ALL=(meta-analysis)) NOT ALL=(animal)) NOT ALL=(huntington)) NOT ALL=(Hiv)) NOT ALL=(schizophrenia)) NOT ALL=(Multiple Sclerosis)) NOT ALL=(Autism)) NOT ALL=(epilepsy)) NOT ALL=(Cerebral palsy)) NOT ALL=(Seizure)) NOT ALL=(infection)) NOT ALL=(injury)) NOT ALL=(Amyotrophic Lateral Sclerosis)) NOT ALL=(Diabetes Mellitus)) NOT ALL=(depression)) NOT ALL=(anxiety)) NOT ALL=(bipolar)) NOT ALL=(psychosis)) NOT ALL=(glaucoma)) NOT ALL=(cell)) NOT ALL=(receptor)) NOT ALL=(protein)) NOT ALL=(serum)) NOT ALL=(lipid)) NOT ALL=(covid)) NOT ALL=(cancer)) NOT ALL=(apnea)) NOT ALL=(case report)) NOT ALL=(glucose)) NOT ALL=(syndrome)) NOT ALL=(stroke)) NOT ALL=(delirium)) NOT ALL=(Atherosclerosis)) NOT ALL=(Damage*)) NOT ALL=(Tumor)

#### Final search string

Search title/abstract: **#1** AND **#2** AND **#3** = **#4**

After exclusion: **#4** NOT **#5**

### Search strategy for PsycINFO

*Concept #1: Dementia*

Dementia/ OR Alzheimer’s Disease/

*Concept #2: Neuropsychological tests*

cogniti*/ or neuropsychological test*/ or memory

*Concept #3: Prediction, progression, conversion*

predict*/ or diagnos*/ or coversion/ or progress*/ or trajector*/ or detect*/ or Differentiat*/ or distinguish

*Concept #4: Exclusion of irrelevant terms*

limit 4 to (peer reviewed journal and human and english language and adulthood <18+ years> and “300 adulthood <age 18 yrs and older>“)

#### Final search string

**#1** AND **#2** AND **#3** AND **#4**

## Notes

**Competing interests** The authors declare no conflicts of interest.

### Competing Interest Statement

The authors have declared no competing interest.

### Funding Statement

This study did not receive any funding.

